# Pfs230 yields higher malaria transmission-blocking vaccine activity than Pfs25 in humans but not mice

**DOI:** 10.1101/2020.11.19.20234922

**Authors:** Sara A. Healy, Charles Anderson, Bruce J. Swihart, Agnes Mwakingwe, Erin E. Gabriel, Hope Decederfelt, Charlotte V. Hobbs, Kelly M. Rausch, Daming Zhu, Olga Muratova, Raul Herrera, Puthupparampil V. Scaria, Nicholas J. MacDonald, Lynn E. Lambert, Irfan Zaidi, Camila H. Coelho, Jonathan P. Renn, Yimin Wu, David L. Narum, Patrick E. Duffy

## Abstract

**Background:** Vaccines that block human-to-mosquito *Plasmodium* transmission are needed for malaria eradication and clinical trials have targeted zygote antigen Pfs25 for decades. We reported that a Pfs25 protein-protein conjugate vaccine formulated in alum adjuvant induced significant serum functional activity in both US and Malian adults. However, antibody titers declined rapidly, and transmission-reducing activity required four vaccine doses. Functional immunogenicity and durability must be improved before advancing TBV further in clinical development. We hypothesized that the pre-fertilization protein Pfs230 alone or in combination with Pfs25 would improve functional activity.

**Methods:** Transmission-blocking vaccine candidates based on gamete antigen Pfs230 or Pfs25 were conjugated with Exoprotein A, formulated in Alhydrogel^®,^ and administered to mice, rhesus macaques, and humans. Antibody titers were measured by ELISA and transmission-reducing activity was assess by the Standard Membrane Feeding Assay.

**Results:** Pfs25-EPA/Alhydrogel^®^ and Pfs230D1-EPA/Alhydrogel^®^ induced similar serum functional activity in mice, but Pfs230D1-EPA induced significantly greater activity in rhesus monkeys that was enhanced by complement. In U.S. adults, two vaccine doses induced complement-dependent activity in 4 of 5 Pfs230D1-EPA/Alhydrogel® recipients but no significant activity in five Pfs25-EPA recipients, and combination with Pfs25-EPA did not increase activity over Pfs230D1-EPA alone.

**Conclusion:** The complement-dependent functional immunogenicity of Pfs230D1-EPA represents a significant improvement over Pfs25-EPA. The rhesus model is more predictive of the functional human immune response to Pfs230D1 than is the mouse model.

**Trial Registration:** ClinicalTrials.gov NCT02334462

**Funding:** This work was supported by the Intramural Research Program of the National Institute of Allergy and Infectious Diseases, National Institutes of Health.

## Introduction

The world has achieved significant strides in malaria control with roughly half of countries having eliminated the disease, but existing tools failed to achieve elimination despite comprehensive application in African settings (1, 2), and recent progress to reduce malaria cases has stalled globally and been reversed in some areas (3). Vaccines have been essential for elimination of infectious agents like smallpox, polio, and measles by halting their onward transmission, conferring major benefits to human health and economies (4-6). Malaria transmission-blocking vaccines (TBV) were conceived in the 1970’s as a tool to interrupt parasite transmission with antibodies that attack sexual stage parasites in the mosquito vector (7, 8). Monoclonal antibodies to mosquito sexual stage (gamete) parasites were used to identify candidate TBV antigens, including gamete surface proteins P230 and P48/45 first expressed by gametocytes in human blood (9), and zygote surface proteins P25 and P28 expressed only post-fertilization in the mosquito host (10, 11). These antigens are multi-domain cysteine-rich proteins and generally difficult to produce as properly folded recombinant protein. *P. falciparum* P25 (Pfs25) antigen was the first expressed as recombinant protein (12) and has remained the leading TBV candidate for three decades. In preclinical studies, Pfs25 vaccines have induced equal or greater serum activity versus other candidate antigens or antigen combinations (13, 14).

While earlier P25 candidates failed to meet safety or activity criteria to advance in the clinic (15, 16), we recently reported that a Pfs25 protein-protein conjugate vaccine formulated in alum adjuvant induced significant serum functional activity in both US (17) and Malian adults (18). However, few vaccinees developed transmission reducing activity (TRA) >50% or significantly increased their TRA after 2 or 3 doses of Pfs25 vaccine: 2/17 subjects after 2 doses and 2/15 after 3 doses in US adults (17); and no significant activity was seen in vaccinees (versus comparators) after 3 doses in Malian adults (18). In each study, significant functional activity required four vaccine doses, and antibody titers declined rapidly, suggesting functional immunogenicity and durability must be improved before advancing TBV further in clinical development. We hypothesized that the pre-fertilization protein Pfs230 alone or in combination with Pfs25 would improve functional activity.

To assess the contribution of Pfs230 to a TBV, a fragment (Ser_542_ to Gly_736_) encompassing domain 1 of Pfs230 cloned and expressed in *P. pastoris* (Pfs230D1, previously referred to as Pfs230D1M) as described in (19) was chemically conjugated to EPA (20), a non-toxic mutant of exoprotein A from *P. aeruginosa* using methods previously described for development of the Pfs25-EPA vaccine (21). Here, we compare Pfs230D1-EPA to our benchmark TBV (Pfs25-EPA) formulated in alum in three models (mice, nonhuman primates, and humans) and assess their activity in combination.

## Results

To confirm the benefit of conjugation of Pfs230D1, groups of CD-1 mice were immunized twice (0, 28 days) by intramuscular injection with either Pfs230D1 or Pfs230D1-EPA, both formulated in Alhydrogel^®^, which was the clinical formulation. Antibody titers induced by Pfs230D1-EPA were ∼100-fold greater than titers induced by Pfs230D1 monomer (**Fig. S1**). For clinical development planning, we assessed Pfs230D1 and Pfs25 for their relative capacities to induce functional antibodies, to determine if one antigen should be prioritized over the other for clinical testing. Initial dose ranging studies with Pfs230D1-EPA were performed in mice in order to determine appropriate doses to use for future mouse experiments (**Fig. S2**); subsequently all mice were immunized with doses ranging from 0.1 to 3 ¼g per immunization 28 days apart.

Data were collated from multiple experiments using both BALB/c and CD-1 outbred mice with head-to-head comparisons of Pfs25-EPA/Alhydrogel^®^ and Pfs230D1-EPA/Alhydrogel^®^ for functional antibody activity assessed by Standard Membrane Feeding Assay (SMFA) of *P. falciparum* strain NF54 gametocytes to *Anopheles stephensi* mosquitoes (**Fig. 1A**). Parasite Transmission-Reducing Activity (TRA, reduction in mean oocyst count per mosquito versus control antibody) was plotted against ELISA titers for 4 separate experiments, containing 3 immunizing doses in each experiment. There was a titer-dependent positive correlation for both antigens (Pfs230D1, r=0.68 P=0.02; Pfs25, r=0.58 P=0.06), with high TRA achieved at the higher titers with both antigens, but with no difference between Pfs25 and Pfs230D1.

**Fig. 1.**
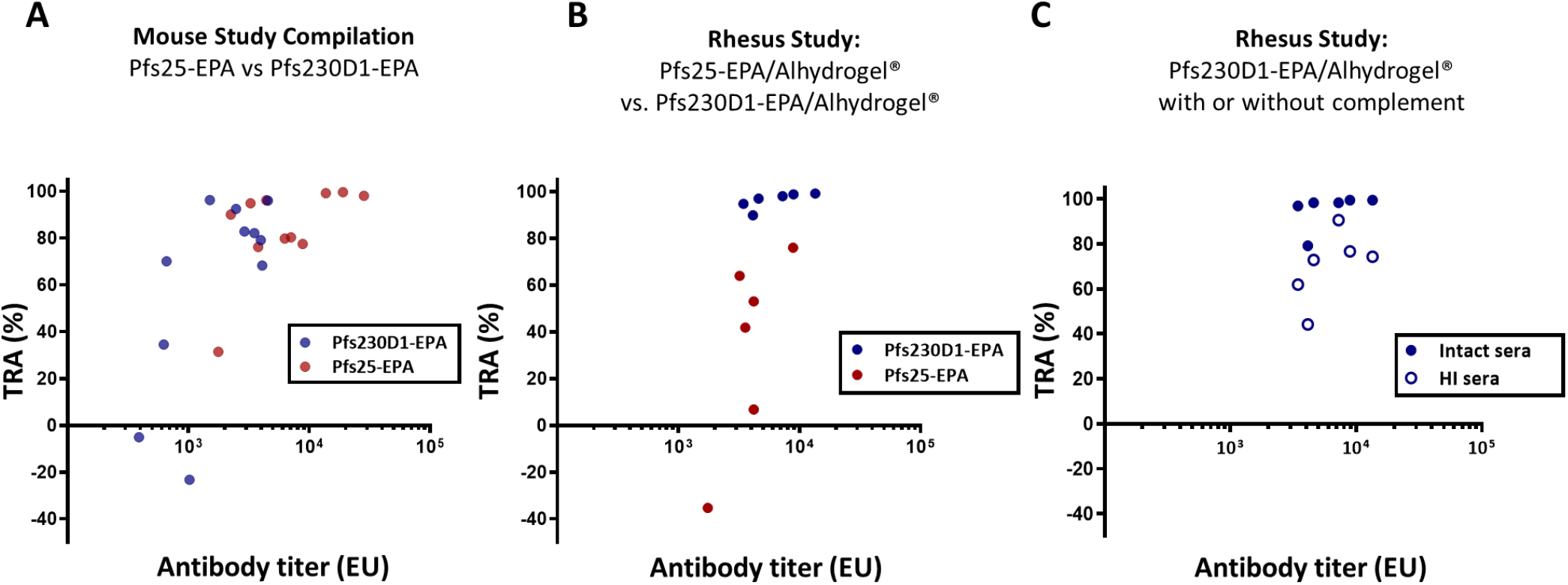
While (A) mouse studies comparing Pfs25-EPA/Alhydrogel® versus Pfs230D1-EPA/Alhydrogel® revealed no difference in immunogenicity or functional activity, (B) rhesus studies suggest Pfs230D1 is superior to Pfs25 and (C) requires active complement. (A) Mouse sera samples collected after immunization with Pfs25-EPA/Alhydrogel® or Pfs230D1-EPA/Alhydrogel® were used in SMFA to measure antibody function. Each data point represents a pool of sera from one immunization group (n=10/grp). The titer shown in the x-axis is the titer in the mosquito feeder after dilution. (B) Rhesus monkey sera samples collected two weeks after the third immunization with Pfs25-EPA/Alhydrogel® or Pfs230D1-EPA/Alhydrogel® were used in SMFA to measure antibody function. Each data point represents an individual animal. The titer shown on the x-axis is the tier in the mosquito feeder after dilution. %TRA is relative to pre-bleed samples from each animal; (C) Sera from the Pfs230D1-EPA/Alhydrogel® group were divided in two for heat inactivation followed by SMFA.

Multiple publications have shown a dependency on active complement for anti-Pfs230 inhibition in preclinical studies (22-24). Therefore, serum samples from BALB/c mice immunized with cGMP Pfs230D1-EPA/Alhydrogel^®^ were tested by SMFA when mixed with intact or heat-inactivated human sera. Again, sera from mice immunized with Pfs230D1-EPA inhibited oocyst development in a dose-dependent manner and was not significantly different from that of Pfs25-EPA sera (**Fig. S3, Table S1**). In the absence of active human complement in a dose titration experiment, antibodies against both antigens blocked equally well.

Since mouse studies did not distinguish one antigen (Pfs25 or Pfs230D1) as superior over the other for functional activity and therefore their prioritization for clinical testing, we hypothesized a non-human primate model may reveal quantitative differences in activity of the two vaccine candidates. We conducted a vaccine study in rhesus macaques using the clinical Alhydrogel® formulation and doses of Pfs230D1-EPA (40 µg Pfs230D1) and Pfs25-EPA (47 µg Pfs25) administered by intramuscular injection on a 0, 2, 6-month schedule, similar to a typical human clinical trial regimen and schedule. Antibody titers were monitored to confirm Pfs25 and Pfs230D1 immunogenicity (**Fig. 1B, Fig. S4A, C**). Sera from peak titers, collected two weeks after the third dose (day 182), were used in SMFA to assess function in the presence of intact human sera (**Fig. 1B, Fig. S4B, D, and Table S2**). Anti-Pfs25 immune sera demonstrated overall modest inhibition of oocyst development, similar to previous preclinical and clinical experience. In contrast, anti-Pfs230D1 had potent inhibitory activity, with all samples reducing oocyst density by greater than 80%, and 3/6 achieving greater than 50% reduction in the prevalence of infected mosquitoes, referred to as Transmission-Blocking Activity (TBA).

To measure the contribution of complement to the activity, serum samples from the Pfs230D1 group were tested again by SMFA, in the presence or absence of complement (**Fig. 1C and Table S3**). Distinct from observations in mice, activity was significantly diminished in the absence of complement (p=0.03, Wilcoxon Matched-Pairs signed rank test for difference between groups). However, even without complement, 5/6 monkeys had greater than 50% TRA, suggesting rhesus react to an epitope that blocks biological function. Thus, the rhesus model indicated that Pfs230D1 was a superior vaccine target, in part due to the activity of complement.

After establishing that Pfs230D1-EPA/Alhydrogel^®^ or Pfs25-EPA/Alhydrogel^®^ were well-tolerated, immunogenic, and induced functional activity in preclinical models, a phase 1 first-in-human clinical trial was initiated to assess safety and to compare Pfs25-EPA/Alhydrogel^®^, Pfs230D1-EPA/Alhydrogel^®^, or a combination of the two, before advancing either candidate to field trials. US adults were vaccinated in a staggered manner for safety. Five subjects per arm received two immunizations (0, 28 days) with Pfs230D1-EPA/Alhydrogel^®^ or Pfs25-EPA/Alhydrogel^®^ or both co-administered in separate arms. Overall, vaccinations with all doses of Pfs25-EPA/Alhydrogel^®^ (16 µg; 47 µg), Pfs230D1-EPA/Alhydrogel^®^ (5 µg; 15 µg; 40 µg), and the combination (Pfs25 + Pfs230D1: 16 µg + 15 µg; 47 µg + 40 µg) were well-tolerated with minimal local and systemic reactogenicity reported and no serious adverse events (**Table S4**). Pfs25-EPA/Alhydrogel^®^ elicited anti-Pfs25 antibodies at the high dose but not at the low dose (**Fig. 2B** compared to **2A**, p=0.03) while titers against Pfs230D1 were not dose-dependent (**Fig. 2C, D, E**). Combining low dose Pfs25-EPA/Alhydrogel^®^ with low dose Pfs230D1-EPA/Alhydrogel^®^ elicited significantly higher anti-Pfs25 titers than low dose Pfs25-EPA/Alhydrogel^®^ alone (**Fig. 2F** compared to **2A**, p=0.01); this effect was not observed with high dose vaccine (**Fig. 2G** compared to **2B**, p=0.12), and there was no potentiating effect of vaccine combinations on anti-Pfs230 titers. Interestingly, anti-Pfs230 titers were measurable even at the lowest Pfs230D1-EPA dose of 5 ug (**Fig 2C**), unlike the Pfs25-EPA vaccine (18), suggestive of better intrinsic immunogenicity.

**Fig. 2.**
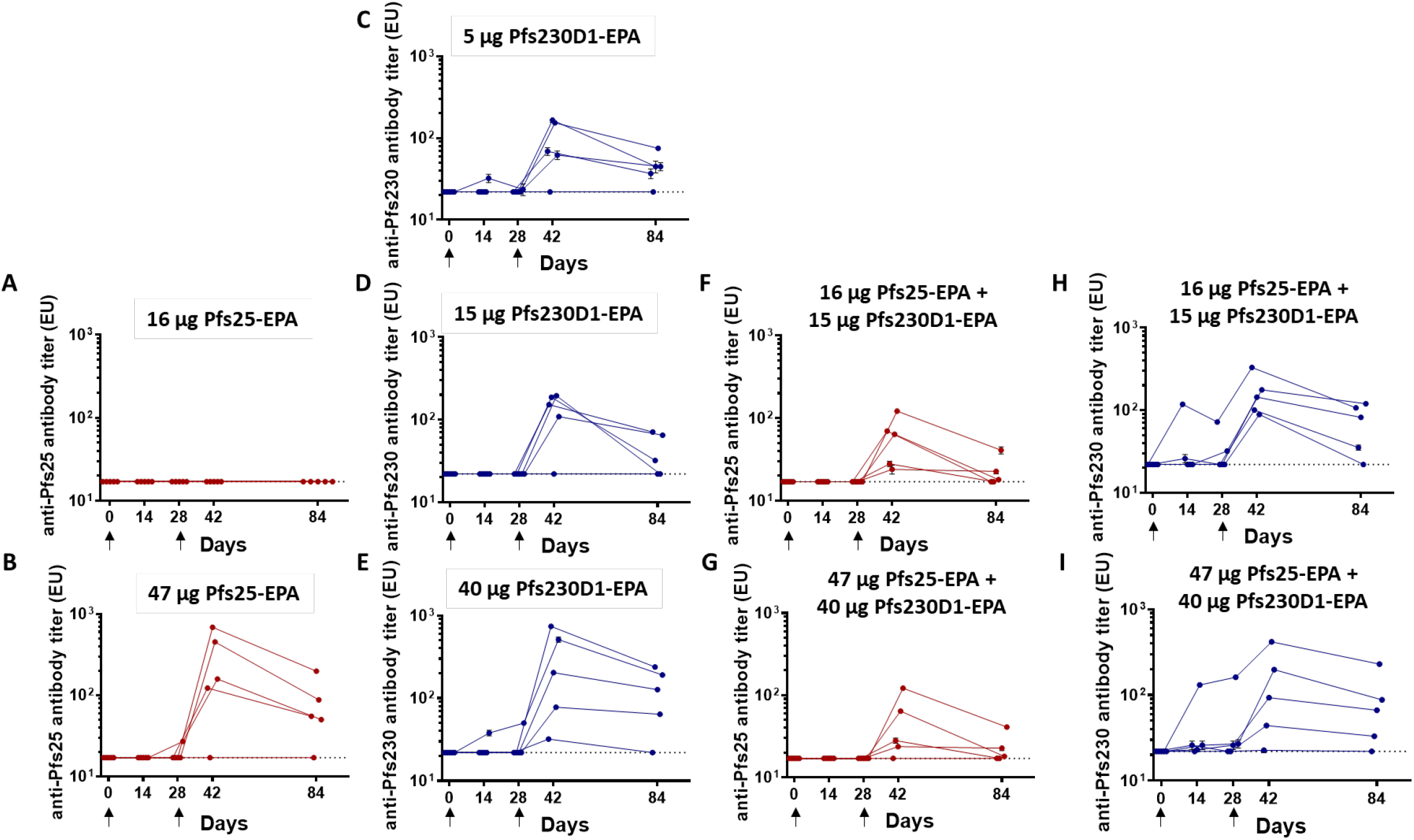
Antibody titers by ELISA in humans. Antibody titers from each vaccine arm. Vaccinations were performed on days 0 and 28 at escalating doses (n=5/arm).

Sera from peak titers two weeks after the second dose in the high dose groups were measured for functional activity (TRA and TBA) by SMFA. In general, TRA are used to compare results from different studies owing to consistency between assays, while TBA varies between assays based on parasite infectivity to mosquitoes (25). Consistent with previous published data (17) and with preclinical studies, two doses of Pfs25-EPA did not induce titers sufficient to reduce transmission (**Fig. 3A and Table S5**). However, aligning with the results seen in rhesus, Pfs230D1 induced high functional activity in humans. TRA was >90% in 2/5 individuals from the Pfs230D1-EPA arm (99%, 98%) and > 50% TRA in 2 others (73%, 62%), versus the Pfs25 group in which 0/5 individuals had TRA>50%, representing a significant improvement of Pfs230D1-EPA over the benchmark Pfs25-EPA candidate (Fishers exact test, p<0.05). In the Pfs25 + Pfs230D1 combination group, one individual had 90% and one had 65% TRA. TRA correlated well with anti-Pfs230D1 titers (r=0.77, p=0.013) but not anti-Pfs25 titers (**Fig 3B-C**), demonstrating that the functional activity is due to the Pfs230D1 vaccine. Titers achieved after 2 doses of Pfs25 vaccine were similar in two previous studies in malaria-naïve US individuals (GM EU=147) (17) and malaria-exposed individuals in Mali (GM EU=93.1) (18) as in this study (GM EU=160.1), when comparing individuals who received the same vaccine dosage. Functional Pfs25 activity was also similar between the prior US study (17) and this study, with few (2/17) or no (0/5) individuals with serum TRA>50%, respectively, after 2 vaccine doses; serum TRA was not measured after vaccine dose 2 in the Mali field trial (18) and so cannot be compared to the current study.

**Fig. 3.**
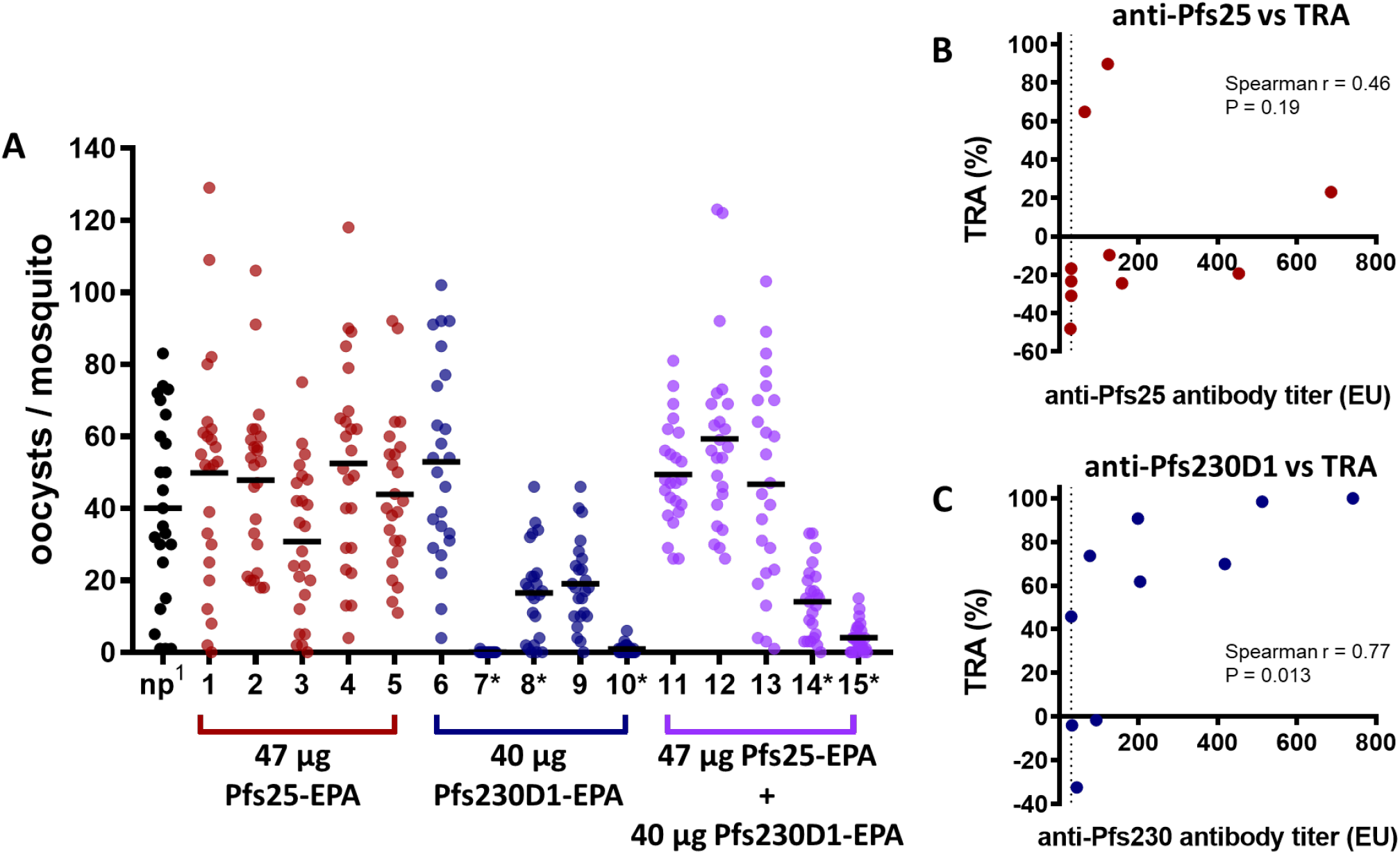
Anti-Pfs230D1 has functional activity after 2 doses in humans. Vaccination induces functional antibodies after 2 doses. Sera from subjects (n=5/grp) were collected 2 weeks after 2nd dose and used for SMFA. Each data point represents the oocyst count from one mosquito. n.p., naïve sera pool. * p<0.05 difference from naïve pool, 15 pairwise Kruskal-Wallis tests with Bonferroni correction for multiple comparisons. Figure data are also represented in **Table S5**.

Again, as seen in the non-human primate model, dependency on complement was confirmed with the Pf230D1 immune sera samples, as heat-inactivation of serum complement significantly reduced (p=0.009, Wilcoxon matched-pairs signed rank test), but did not completely eliminate, functional activity (**Fig. 4A, B** and **Table S6**). Sera from two individuals who received the vaccine combination appeared to increase parasite transmission to mosquitoes after heat-inactivation (**Fig. 4D**); enhancement of *P. vivax* transmission has also been observed with naturally acquired antibodies or monoclonal antibodies used at low doses in membrane feeding assays (26). Naturally acquired serum transmission-enhancing activity has been observed against both *P. vivax* and *P. falciparum* (26-28), and this enhancing activity has been most apparent in heat-inactivated sera (27). While this enhancing activity has not previously been associated to Pfs230 antibodies in the absence of complement, our results echo these earlier findings.

**Fig. 4.**
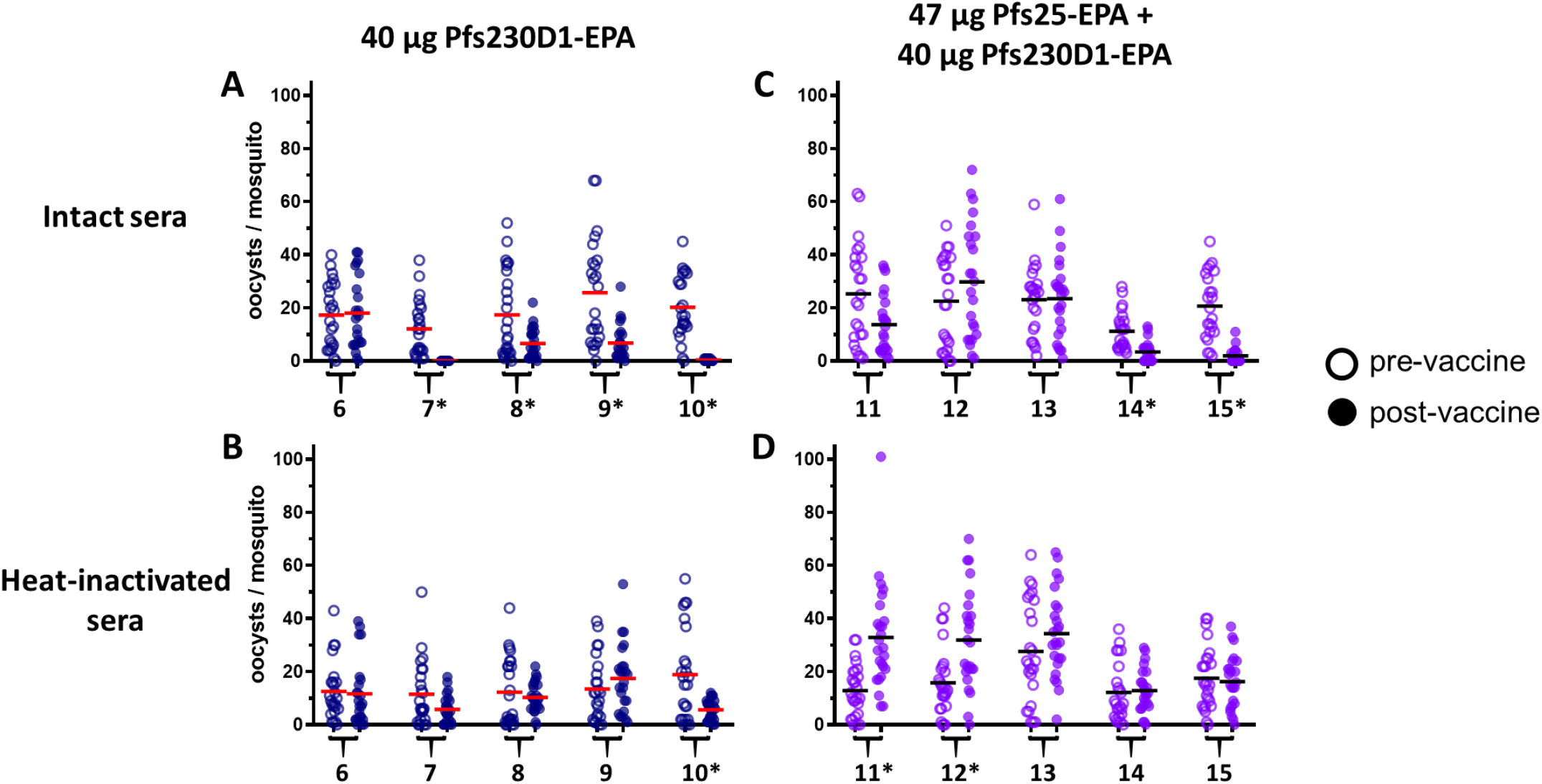
Functional activity of anti-Pfs230 requires active complement in humans. Pfs230D1 requires complement for activity. Sera from subjects vaccinated with Pfs230D1-EPA/Alhydrogel or Pfs230D1-EPA+Pfs25-EPA/Alhydrogel were divided in two for complement inactivation and used for SMFA. Each data point represents the oocyst count from one mosquito. *p<0.05 difference between post-vaccine and pre-bleed sera from individuals, Wilcoxon matched pairs signed-rank test with Bonferroni adjustment. Figure data are also represented in **Table S6**.

## Discussion

A transmission blocking vaccine could play a pivotal and unique role in malaria elimination efforts, by inducing durable immune responses that interrupt human-to-mosquito malaria transmission over an extended period of time. In our earlier clinical trials, the leading TBV candidate Pfs25 induced functional activity at peak antibody titers, but the activity was lost rapidly as titers decreased (17, 18). Here, we sought to increase TBV activity and durability by combining the post-fertilization antigen Pfs25 with the pre-fertilization antigen Pfs230D1. We found that Pfs230D1 was significantly more potent than Pfs25 as a vaccine, and the addition of Pfs25 to Pfs230D1 did not appear to improve on this activity. The superior activity of Pfs230D1 over Pfs25 was predicted by preclinical studies in rhesus but not in mice and was largely explained by complement-dependent activity seen with monkey and human but not rodent antibody. Pfs230 or Pfs230 combination vaccines warrant further evaluation in field studies to assess their potential as a tool for malaria elimination as do adjuvants that increase levels and durability of functional antibody (29), particularly complement-fixing antibody isotypes.

TBV candidate antigens including Pfs230 have been challenging to prepare as properly folded proteins, owing to their cysteine-rich sequence and highly folded structure. Importantly, functional antibodies against these antigens commonly target conformation-dependent epitopes (19). The Pfs230 extracellular fragment includes 14 domains with 6-cys structure, a motif that has been particularly difficult to reproduce as recombinant antigen. Earlier studies showed that a Pfs230 polypeptide (Pfs230-C, amino acids 443-1132) encompassing an upstream region, domain 1 through 3, and 9 amino acids in domain 4 (30) or a polypeptide (Pfs230D1-2) encompassing an upstream region, domain 1 through 2 (19), induces functional antibodies in animal studies Here, we used *Pichia* expression system to generate properly folded Pfs230 domain 1 which induced more potent antibodies in rabbits compared to the Pfs230D1-D2 polypeptide in the *ex vivo* SMFA (19), after prior success expressing full-length extracellular Pfs25 in *Pichia* (31) that induced functional antibodies in human studies (17). Our preclinical studies consistently demonstrated *Pichia*-expressed Pfs230D1 reacted to conformation-dependent functional monoclonal antibodies (19) and induced functional IgG in immunized animals.

Recent studies in mice suggested that Pfs25 immunogens were similar (13) or superior (14) to Pfs230 immunogens for inducing functional antibodies that block parasite transmission. In mice, we similarly found no significant differences in the ability of Pfs25 and Pfs230D1 recombinant immunogens prepared as conjugate nanoparticle vaccines to induce antibody activity in mice. However, our studies in monkeys suggested that the Pfs230D1 immunogen was significantly more potent for inducing serum functional activity. The difference between models can be explained at least partly by the complement-dependent nature of Pfs230-induced activity, which was pronounced in assays with monkey but not mouse antibody. Early studies using mouse monoclonal antibodies raised against native Pfs230 on gametes revealed functional antibodies restricted to IgG2a and IgG2b subclasses, with complement-mediated lysis being the putative mechanism of transmission blockage (22-24) with the exception of the murine mAb 4F12, isotype IgG1 which has been crystallized in complex with Pfs230D1 (32) and effectively blocks transmission independent of complement (19).

We examined antibody subclasses and isotypes to assess the role of complement-fixing antibody. In a previous study, we showed that mice immunized with Pfs230D1 showed a dominant IgG1 isotype response (33) which is non-complement-fixing in mice. In the present study, we measured Pfs230D1-specific antibody isotypes in sera from rhesus macaques that received either Pfs230D1-EPA/Alhydrogel® alone or Pfs25-EPA/Alhydrogel® + Pfs230D1-EPA/Alhydrogel® combination, two weeks after the second and third vaccinations (**Fig. S5**); and from human vaccinees 2 weeks after the second vaccination (**Fig. S6)**. In both rhesus and humans, we observed that IgG1, which has the highest complement-fixing ability, was the dominant isotype induced, while the non-complement fixing IgG2 and IgG4 were significantly lower. Two human vaccinees produced appreciable amounts of IgG3 which is also complement-fixing. Pfs230-specific IgM, which also have complement-fixing properties, was readily detected in 4 of the 5 vaccinees but they were consistently lower than the IgG1 isotype in all samples. We surveyed Day 84 sera for five subjects that had TRA on Day 56, and significant functional activity persisted for 4/5 subjects (**Fig. S7A**). We compared IgG and IgM purified from Day 84 sera of two subjects (Subjects #7 and #10) in SMFA, and oocyst counts were significantly lower in the presence of IgG than IgM (for Subject #7, p = 0.042; for Subject #10, p = 0.006; for both subjects combined, p = 0.001, **Fig. S7B**). Both serum and IgG from Subject #7 were tested for membrane attack complex (MAC) formation on gametes, showing MAC formation in assays using intact but not heat-inactivated sera (**Fig. S7C, D, E, F**). These results are consistent with recent evidence that human monoclonal antibodies we generated from Pfs230D1 vaccinees also induce MAC formation on gametes in the presence of intact but not heat-inactivated sera (34).

Of note, mouse polyclonal sera induced by vaccination with an alum-based adjuvant conferred activity in the absence of the complement pathway. Additionally, rhesus immune sera significantly reduced transmission in the absence of complement, demonstrating other complement-independent mechanisms, such as neutralization. Indeed, in the clinical trial, function in one subject was not completely ablated in the absence of complement (**Fig. 4B**). We are currently exploring mechanisms by which rodent mAb 4F12 might neutralize gametes in the absence of complement activity.

Our initial goal for the human studies was to examine whether a combination of Pfs25 with the pre-fertilization antigen Pfs230D1 was safe and might induce more potent and longer-lived activity than Pfs25 alone. In previous rodent studies, a combination of yeast-expressed Pfs25 and Pfs28 induced greater activity than either antigen alone (35), but combinations of Pfs25 and Pfs28 or of Pfs25 and Pfs230-C delivered as virus-vectored vaccines did not yield additional activity (14). Ultimately, each antigen combination must be tested empirically for positive or negative interference in humans. Here, we observed that Pfs230D1 alone formulated in Alhydrogel® (which has relatively modest immunopotentiating activity) could induce functional serum activity after only two doses in some individuals, suggesting vaccine activity could be enhanced by increasing the number of doses or by alternative adjuvants. Further, the addition of Pfs25 to Pfs230D1 during immunization did not provide additional activity. We are currently evaluating alternative adjuvants for their ability to enhance TBV activity, by increasing antibody titers and durability particularly for complement-fixing IgG subclasses.

Taken together, the data show two conclusions: 1) Pfs230D1 is a superior transmission-blocking antigen to Pfs25, and 2) the rhesus model is more predictive of the functional human immune response to Pfs230D1 than is the mouse model. The results from these studies yield valuable information for future studies to understand immune responses to Pfs230D1 and how improvements in vaccine development may lead to a licensed TBV.

## Methods

### Animal Studies

Five to 8-week-old naïve, female BALB/c or CD-1 mice were purchased from Taconic Laboratories (Hudson, NY) and maintained at a facility at the NIH. Immunizations were performed by either intraperitoneal or intramuscular (i.m.) injection in the anterior tibialis in a volume of 50 µL using a standard day 0 and day 28 regimen. *Macaca mulatta* (rhesus) were randomized by age, sex, and weight, and were maintained in an AAALAC accredited NIAID facility. Vaccinations were performed on days 0, 56, and 168 by intramuscular injection in a volume of 0.6 mL (Pfs25-EPA/Alhydrogel) or 0.8 mL Pfs230D1-EPA/Alhydrogel) in the leg, alternating legs for boosting injections.

### Clinical Study Procedures

The clinical study was designed as an open label, dose escalation study to examine the safety and immunogenicity of Pfs230D1-EPA/Alhydrogel^®^ and Pfs25M-EPA/Alhydrogel^®^ alone or co-administered. The initial open-label dose-escalating, two-dose regimen (0, 1 month; n=5/group) was performed in the US prior to a larger double-blinded study conducted in Mali. Participants were sequentially enrolled in the following manner. No blinding or placebo arms were implemented. No randomization occurred.

### ELISA

Immulon 4 HBX flat bottom microtiter plates (Dynex Technologies) ELISA plates were coated with 1 μg/ml of antigen in a volume of 100 μL per well in carbonate coating buffer (pH 9.6) overnight at 4°C. After blocking in 5% skim milk in TBS blocking buffer in a volume of 320 μL per well for 2 hrs, samples were serially diluted in TBS/5% milk and plated in triplicate in a volume of 100 μL per well and incubated at room temperature for 2 hours. Plates were washed 4 times and alkaline phosphatase labeled goat anti-mouse IgG (H+L), goat anti-human IgG (H+L), or goat anti-monkey phosphatase labeled secondary antibody (Seracare Life Sciences) was added in a volume of 100 μL per well and incubated at room temperature for 2 hours. After washing 4 times, dissolved phosphatase substrate tablets (Sigma) were added in a volume of 100 μL per well and plates were incubated for 20 minutes before optical densities (OD) were measured with a Spectramax 340PC (Molecular Devices). Each ELISA plate contained an internal serum standard from which a four-parameter curve was calculated with Softmax software. According to laboratory SOP, any samples for which ELISA results from triplicate wells exceeded a pre-specified CV were repeated. ELISA Units were assigned to test samples based on the sera dilution that gave an OD of 1.0, adjusted to the internal standard. For the Pfs230 isotyping assays, similar procedures were followed and the list of detecting antibodies utilized are listed in Supplementary table S7.

### Transmission Blocking and Reducing Activity

Transmission blocking activity (TBA, reduction in infection prevalence) and transmission reducing activity (TRA, reduction in infection intensity) of the sera were tested by an *ex vivo* standard membrane feeding assay (SMFA) as described previously *(13)*. Briefly, an *in vitro* 14-16 day old gametocyte culture of *P. falciparum* (NF54 line) was evaluated for stage V gametocytes (>0.5%) and the presence of exflagellation centers observed at 400X magnification (>1 per field). The culture was diluted with washed O+ red blood cells (RBCs) from a malaria naïve donor (Interstate Blood Bank, Memphis, Tennessee) to achieve 0.12% ± 0.05% concentration of Stage V gametocytes. For each sample, 100 µL of the pelleted diluted culture (100% hematocrit) was mixed with 160 µL of test serum. For human sera samples, 160 ¼L was used neat; for rhesus sera samples, 60 ¼L of test sera was mixed with 100 ¼L of a pool of naïve human AB^+^ sera; for mouse sera samples, 20 or 30 ¼L of test sera was mixed with 130 ¼L of pooled naïve sera. All samples were immediately fed to pre-starved (∼24 hours) 3-8 day old *Anopheles stephensi* (Nijmegen strain) mosquitoes through a Parafilm^®^ membrane stretched across a glass mosquito feeder kept warm by a circulating water membrane at 40°C. Test sera were not heat-inactivated. Post feeding, mosquitoes were maintained for 8 days at 27°C and 80% humidity conditions to allow for the development of parasites. Infectivity was measured by dissecting at least 20 gravid mosquitoes per sample, staining the midguts with 0.05% mercurochrome solution in water for 20 minutes and counting the number of oocysts on each midgut. The feeding experiment was not analyzed unless the average oocyst count in the assay control mosquitoes (at least 20 dissected mosquitoes fed with naïve heat-inactivated serum) was more than four. The TBA and TRA are calculated by the following formulas:

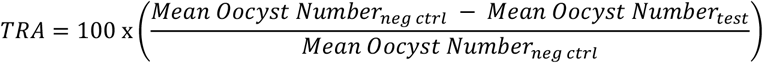

and

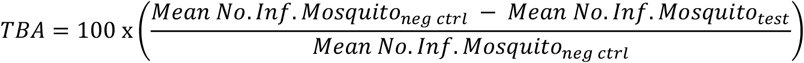

where the negative control (*neg ctrl*) feed used pooled pre-vaccination sera from all subjects. Each sample from monkeys or humans that received Pfs230 or Pfs230 + Pfs25 vaccine was tested in two independent feeding experiments and these two TRA values were averaged to obtain a single subject level TRA for a given time point.

### Immunofluorescence assay to assess the deposition of the membrane attack complex (MAC)

Formation of membrane attack complex (MAC) and further activation of the complement system were assessed using Immunofluorescence of live female gamete parasites. Briefly, 5 mL of a Plasmodium falciparum NF4 gametocyte stage V culture were centrifuged at 2,000 rpm for 5 minutes and added to an exflagellation medium containing 900 µL of RPMI, 100 µL of PBS and 1 µL of xanthurenic acid, then left for 1.5 hours at room temperature. Cells were resuspended in 5 mL of RPMI and applied to a 15 mL Nycodenz gradient (16%, 11% and 6%), then centrifuged at 7,000 x g for 30 minutes. Parasites located in the interface between 6% and 11% were collected into 50 mL of RPMI and spun down at 2,000 x g for 10 minutes. Parasites were then incubated with serum or 100 µg/mL of total IgG previously purified from subject 30G and diluted in PBS. IgG1 isotype control with heavy and kappa chains was purchased from Creative Biolabs. The suspension was incubated at 37° C for 45 minutes with either 50 μL of intact sera or sera heat-inactivated at 56°C, both obtained from US healthy donor. Subject serum was used directly without supplementation. During the incubation the tubes were gently mixed every 10 minutes to facilitate C5b-9 and C5b-8 deposition on cells. Two mL of cold PBS were used to stop the reaction and to wash the cells. Suspension was centrifuged at 500 x g for 5 minutes and the pellet was then incubated with 10ug/mL of the mAb anti-C5b-9 + C5b-8 (Abcam, ref ab66768) for 2 hours on ice. Cells were washed with PBS, centrifuged at 500 x g for 5 minutes and stained with Hoechst 33342 Solution (Thermo Fisher) diluted at 1:20,000 for 8 minutes and further washed with PBS. Cells were kept in parasite culture media until the imaging was performed in a TCS SP8 MP microscope (Leica, Wetzlar, Germany) at 37°C. Quantification analyses was performed assessing the mean fluorescence per nuclei stained.

### Statistics

All statistical tests were performed using Prism v7.0 by GraphPad Software, Inc. Pearson’s correlation coefficient for the pairs of log-titer and TRA stratified by titer type were conducted for Figure 1A. Figure 2, Day 42 values of panel A vs F, B vs G, and A vs B were conducted by separate Wilcoxon rank sum tests with continuity correction. Figure 2 panels C vs D vs E were compared with a Kruskal-Wallis rank sum test. Figure 3A showcases 15 Kruskal Wallis tests (each animal compared with the naïve-pool as a control) with a Dunn-Bonferroni adjustment. Spearman’s coefficient and exact p-values were calculated for both Figure 3B and Figure 3C. Figures 4A-D showcase Wilcoxon matched pairs signed-rank tests; each panel had 5 pairwise tests performed. Asterisks indicate p < 0.05/5 = 0.01. Figure S1 reports exact p-values for each of 3 Mann-Whitney tests. Figure S3 (and Table S1) displays 6 Kruskal Wallis tests performed for intact sera where each comparison involves the control and the p-values are Dunn-Bonferroni corrected. The same procedure was repeated for the Hi-Sera group with a different control in Figure S3. Figure S4 (and Table S2) display the results for 12 pre-vac vs post-vac Wilcoxon matched-pairs signed rank tests. Table S3 contains the results of 6 intact vs heat-inactivated Wilcoxon matched-pairs signed rank tests.

### Study Approval

#### Animal Studies

All animal procedures were performed according to protocols approved by the NIAID and NIH Animal Care and Use Committee. All procedures were in accordance with the Guide for the Care and Use of Laboratory Animal Reports NIH 85-23.

#### Human Ethics Statement

This open label phase 1 trial was performed at the National Institutes of Health (NIH) Clinical Trial Center in Bethesda, MD. The study was conducted under an investigational new drug application (IND) with the US Food and Drug Administration (#16251). The protocol was approved by the Institutional Review Board (IRB) of the National Institute of Allergy and Infectious Diseases (NIAID) and under trial investigation number at ClinicalTrials.gov (NCT02334462). All participants gave written informed consent in order to participate in the study.

## Data Availability

All data are available in the main text or the supplementary materials. All data, code, and materials used in the analysis are available by request to any researcher for purposes of reproducing or extending the analysis.

## Author contributions

SAH, CA, EEG, KMR, YW, PED designed and conceptualized the study. KMR, DZ, RH, PVS, NJM, LEL, CHC, JPR, YW, DLN conducted the experiments. SAH, CA, AM, HD, CVH, OM, IZ conducted clinical investigation. CA, BJS, IZ analyzed the results. CA, BJS, EEG curated the data. SAH and CA visualized data and wrote the manuscript; all authors reviewed and edited the manuscript. SAH and PED supervised the study, and PED supervised all teams.

## Acknowledgments

We are grateful to the following at LMIV: the Clinical Trials Team which includes Regina White, Alemush Imeru, and Kattie Khadar; the Data Manager Rathy Mohan; the Project Manager Sharon Wong-Madden. We thank Vu Nguyen, Richard Shimp, Jr., and Karine Reiter for oversight of the production and release of bulk conjugated vaccine. We also thank Holly McClellan and Weili Dai for their assistance in vaccine quality control evaluation. We are grateful for the work by Emily Kelnhofer and Ashley McCormack on the SMFA results; by Joan Aebig, Andrew Orcutt, and Sarah Brockley for ELISA data; by Nada Alani for isotyping ELISAs; and by Matthew Cowles for antibody purification. Thank you to NIH Clinical Center Outpatient Clinic 8 staff, in particular Karen Evans and Pam Stoll, and to NIH Clinical Center Pharmacy staff. Also, our appreciation to the extensive work completed by the LMIV animal team including Sachy Orr-Gonzalez, Brandi Butler, and Tarik Ouahes. In addition, we thank J. Patrick Gorres for assisting with writing, editing, and visualization.

We thank the CRIMSON team at NIAID for providing assistance in the development and finalization of the case report forms and database. We thank OCRPRO for providing external monitoring services for this study and ongoing support and guidance to our NIAID programs, specifically Erin Rudzinski, Lisa Giebeig, Shelly Simpson, and Susan Vogel.

## Supplementary Materials

Human Study Objectives and Design

Figures S1-S7

Tables S1-S7

