## Supplementary Material for "Pfs230 yields higher malaria transmission-blocking vaccine activity than Pfs25 in humans but not mice"

**This PDF file includes:**

**Human Study Objectives and Design**
**Supplementary table S1**
**Supplementary Figures S1-S7**
**Supplementary Tables S1-S7**

### Human Study Objectives and Design

Human Study Objective

The primary objective of the study was to assess the safety and immunogenicity of Pfs30D1-EPA/Alhydrogel® and Pfs25M-EPA/Alhydrogel® given alone or in combination by co-administration in healthy, malaria naïve US adults. Secondary objectives were to determine functional antibody responses to the Pfs25 and Pfs230 proteins as measured by standard membrane feeding assays (SMFA).

Participants

Non-pregnant, healthy malaria-naive US adults age 18-50 years old were recruited from the Bethesda, Maryland area and were screened for the absence of significant medical conditions. Exclusion criteria included prior history of malaria infection in last 10 years, prior travel to a malaria transmission area in the last 5 years or planned travel during the course of the study, and previous receipt of an investigational malaria vaccine in the last 5 years. Participants were also required to be negative for human immunodeficiency virus, hepatitis B, and hepatitis C, as well as to have clinically normal hematological and biochemistry values.

Interventions

PpPfs25M is a *Pichia*-expressed recombinant Pfs25 with a molecular mass of 18,713 Daltons in an oxidized state. EcEPA is an *E. coli*-expressed recombinant protein with molecular mass of 66,975 Daltons. The Pfs25M-EPA conjugate was produced by reaction between thiolated PpPfs25M and maleimide-activated EcEPA, followed by purification using size-exclusion chromatography. Pfs25M-EPA was subsequently formulated with Alhydrogel®, an aluminum hydroxide gel (Frederikssund, Denmark) used extensively as an adjuvant in licensed human vaccines. The Pfs25M-EPA/Alhydrogel® vaccine was provided as a single-use vial. A 0.2-mL volume is administered for delivery of 16 µg conjugated Pfs25M, 16 µg conjugated EPA, and 320 µg Alhydrogel®. A 0.6-mL volume is administered for delivery of 47 µg conjugated Pfs25M, 47 µg conjugated EPA, and 960 µg Alhydrogel®. Of note, Pfs25M differed from Pfs25H tested in earlier clinical trials *(16, 17)* by elimination of the His-tag previously used for purification.

PpPfs230D1 is a *Pichia*-expressed recombinant a sub-segment ($S_{542}$-$G_{736}$) of Pfs230 with a molecular mass of 21,850 Daltons in an oxidized state. The Pfs230D1-EPA conjugate was produced by reaction between thiolated PpPfs230D1 and maleimide-activated EcEPA, followed by purification using size-exclusion chromatography. The Pfs230D1-EPA/Alhydrogel® vaccine was provided as a single-use vial. A 0.1-mL volume is administered for delivery of 5 µg conjugated Pfs230D1, 4.9 µg conjugated EPA, and 160 µg Alhydrogel®. A 0.3-mL volume is administered for delivery of 15 µg conjugated Pfs230D1, 14.7 µg conjugated EPA, and 480 µg Alhydrogel®. A 0.8-mL volume is administered for delivery of 40 µg conjugated Pfs230D1, 39.2 µg conjugated EPA, and 1280 µg Alhydrogel®.

The Pfs25M, Pfs230D1, the EcEPA, the Pfs25-EPA conjugates, the Pfs230D1-EPA conjugates and the final Pfs25M-EPA/Alhydrogel® and Pfs230D1-EPA/Alhydrogel® vaccines were manufactured in cGMP compliance at the Walter Reed Army Institute of Research Bioproduction Facility. The biochemical and biophysical stabilities, including recognition by confirmation-sensitive, transmission blocking monoclonal antibodies, of the conjugate Bulk Drug Substances (Pfs25M-EPA, Pfs230D1-EPA) and the Final Vialed Products (Drug Products Pfs25M-EPA/Alhydrogel® and Pfs230D1-EPA/Alhydrogel®) were each evaluated annually. The potency of both final vaccines (Pfs25M-EPA/Alhydrogel® and Pfs230D1-EPA/Alhydrogel®) were monitored semiannually during the trial until after the last vaccination. All results indicated the conjugates and formulated vaccines were stable and were in compliance with the preset specifications.

Vaccines were administered by intramuscular injection into the deltoid muscle. Arms were alternated with successive vaccinations if a single vaccination was given. If simultaneous vaccinations were administered, each vaccine was delivered separately in alternate arms. Shortly before vaccination, a study pharmacist withdrew the appropriate volume for the dose each participant was to receive. For Pfs25M, an injection volume of 0.2 mL (Groups 1a and 3a) delivered 16 µg Pfs25M (i.e., conjugates comprised of 16 µg Pfs25M, and 16 µg EPA, and 320 µg Alhydrogel®) and an injection volume of 0.6 mL (Groups 1b and 3b) delivered 47 µg Pfs25M (conjugates comprised of 47 µg Pfs25M, 47 µg EPA, and 960 µg Alhydrogel®). For Pfs230D1, an injection volume of 0.1 mL (Group 2a) delivered 5 µg Pfs230D1 (conjugates comprised of 5 µg Pfs230D1, 4.9 µg EPA, and 160 µg Alhydrogel®), an injection volume of 0.3 mL (Groups 2b and 3a) delivered 15 µg Pfs230D1 (conjugates comprised of 15 µg Pfs230D1, 14.7 µg EPA, and 480 µg Alhydrogel®) and an injection volume of 0.8 mL (Groups 2c and 3b) delivered 40 µg Pfs230D1 (conjugates comprised of 40 µg Pfs230D1, 39.2 µg EPA, and 1280 µg Alhydrogel®). The sample sizes for all arms were for safety.

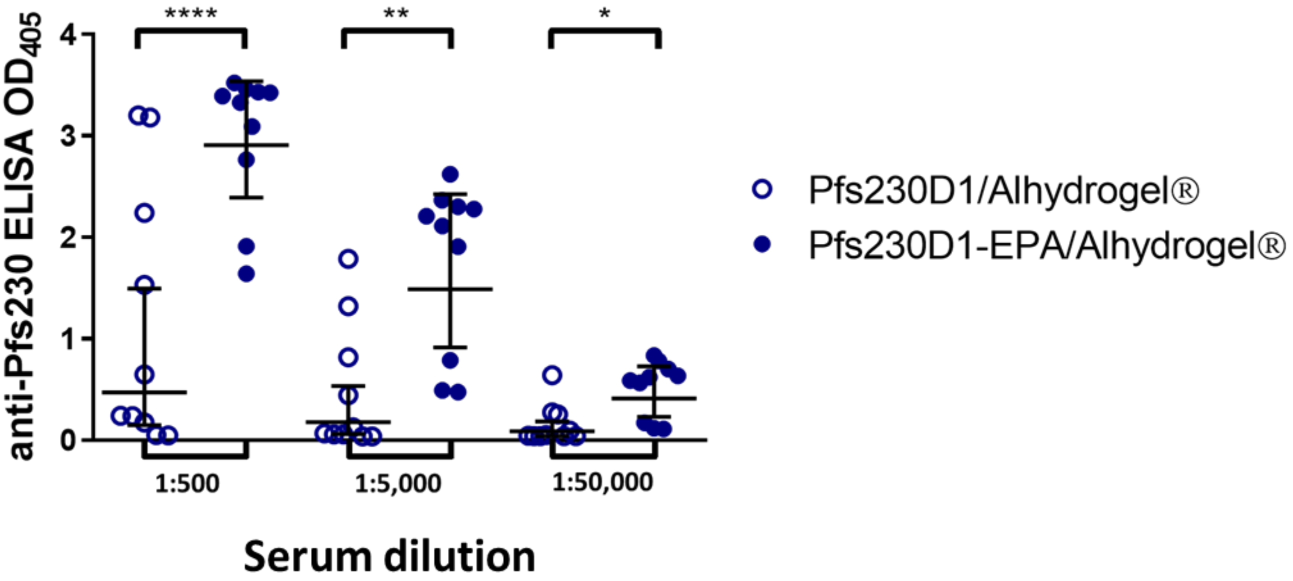

**Fig. S1. EPA enhances immunogenicity and functional activity of Pfs230D1/Alhydrogel® in mice.** CD-1 mice (n=10/group) were immunized i.m. with 0.1 µg of monomeric or conjugated Pfs230D1 on days 0 and 28. Sera were collected on day 42 for ELISA measurements of IgG against Pfs230D1. Values are ODs at the indicated dilution of sera. ***p=0.0015; **p=0.001; *p=0.0038 by Mann Whitney test.

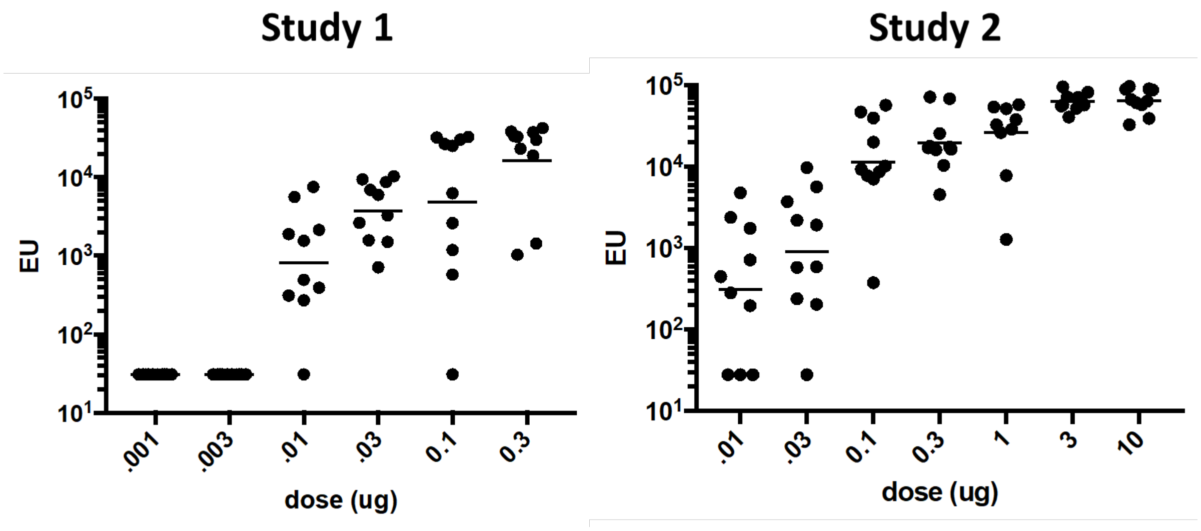

**Fig. S2. Dose-ranging studies of Pfs230D1-EPA/Alhydrogel® in mice.** BALB/c mice (n=10/group) were immunized intraperitoneally with the indicated doses of conjugated Pfs230D1 on days 0 and 21. Sera were collected on day 35 for ELISA measurements of IgG against Pfs230D1. EU = ELISA units.

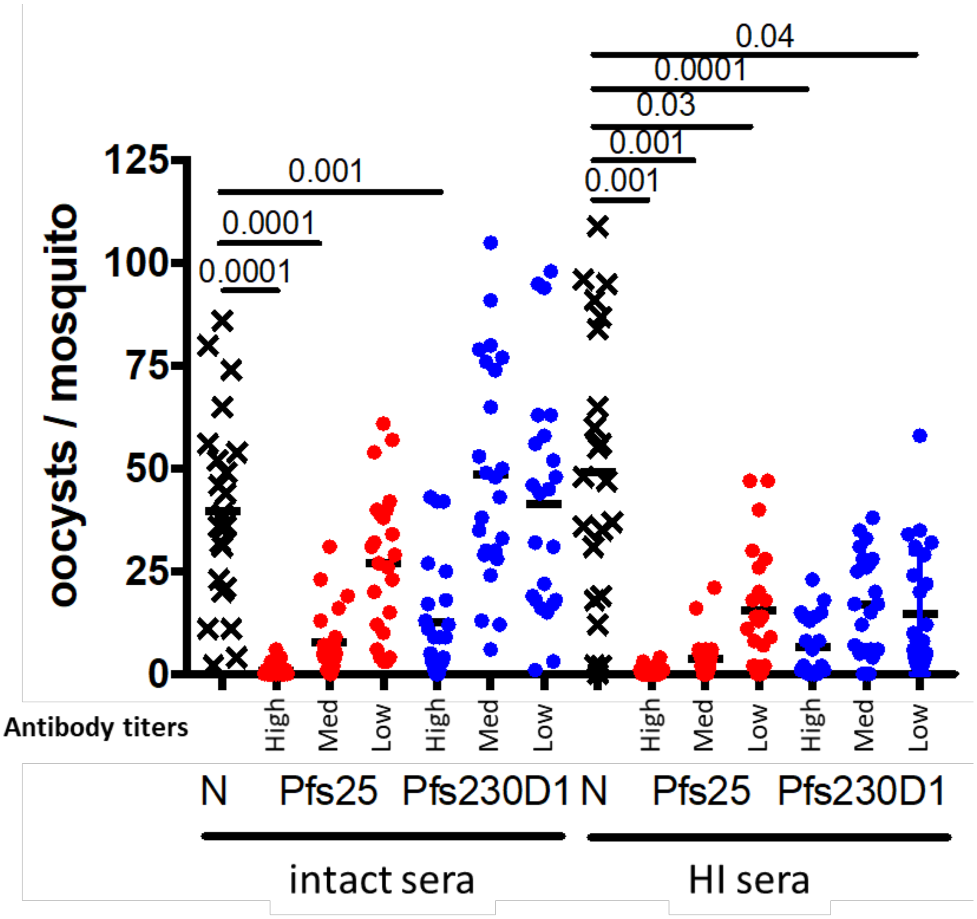

**Fig. S3. SMFA on mouse antisera revealed no superiority in functional activity of Pfs230D1-EPA/Alhydrogel® over Pfs25-EPA/Alhydrogel®, in the presence or absence of complement**

BALB/c mouse sera were collected 2 weeks after immunization with Pfs25-EPA/Alhydrogel® or Pfs230D1-EPA/Alhydrogel® to measure antibody function by SMFA. Sera from each group (n=10/group) were pooled and divided in two for dilution with intact (thus with complement) or heat-inactivated human serum (thus without complement). Three dilutions of each sample were prepared resulting in High (1:8 serum dilution), Medium (Med, 1:32), and Low (1:128) antibody titer groups indicated on the X-axis. **Table S1** provides exact titers for each group. P values are the results from Kruskal-Wallis tests with Bonferroni's correction for multiple comparisons to the naïve control (N). The average number of oocysts in mosquitoes fed with control human AB+ sera was 61. Figure data are also represented in **Table S1**.

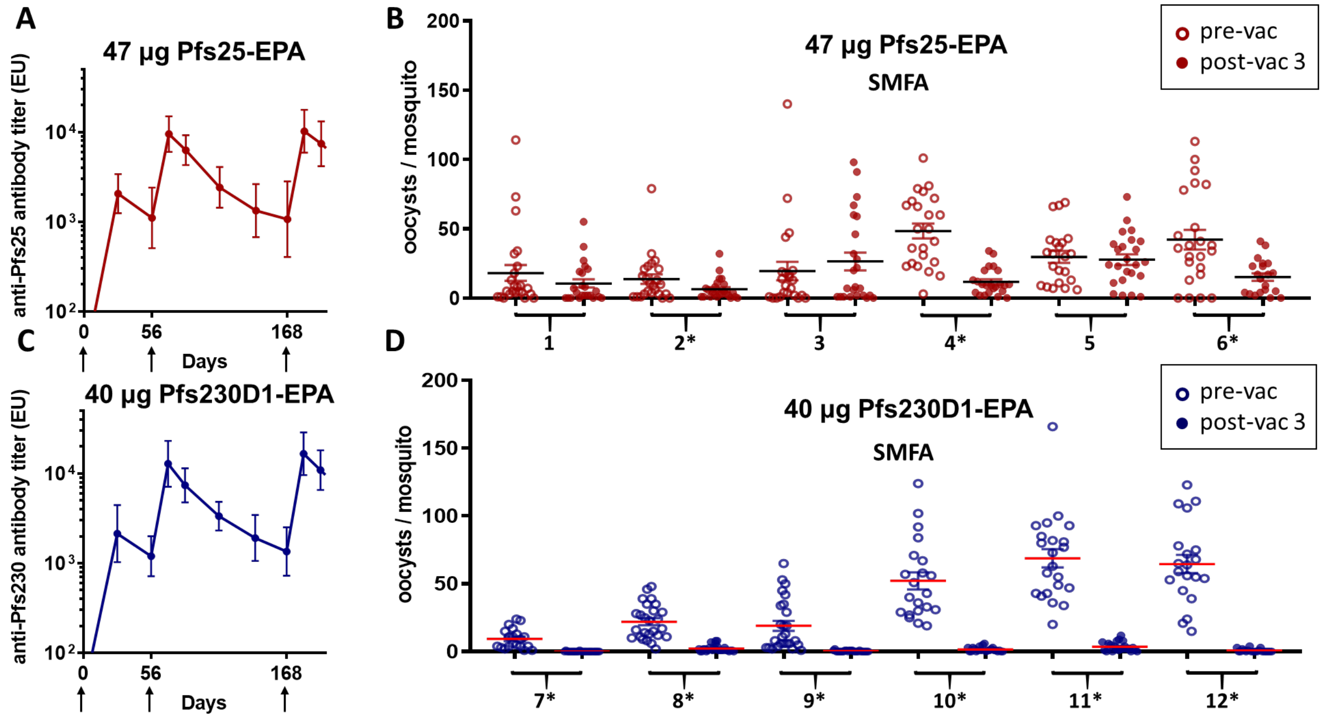

**Fig. S4. Immunogenicity and functional activity of Pfs25-EPA/Alhydrogel® versus Pfs230D1-EPA/Alhydrogel® in rhesus macaques.**

Rhesus monkeys were immunized at 0, 2, and 6 months with Pfs25-EPA/Alhydrogel® or Pfs230D1-EPA/Alhydrogel®. (A) and (C) show antibody titers over time (geometric mean with 95% CI). (B) and (D) show SMFA results from individual rhesus sera samples collected 2 weeks after the third vaccine dose; oocyst counts in negative controls (human AB+ sera) ranged from 16-64. 60 µL of serum from each animal was diluted with 100 µL of $AB^{+}$ human sera, mixed with 100 µL gametocyte culture, and fed to mosquitos. Oocysts were measured 8 days later. Each post-vaccination sample was tested against the pre-vaccine sample, and samples from both vaccine groups were tested side-by-side. Each data point in (B) and (D) represents the oocyst burden from one mosquito. Figure data are also represented in **Table S2**.

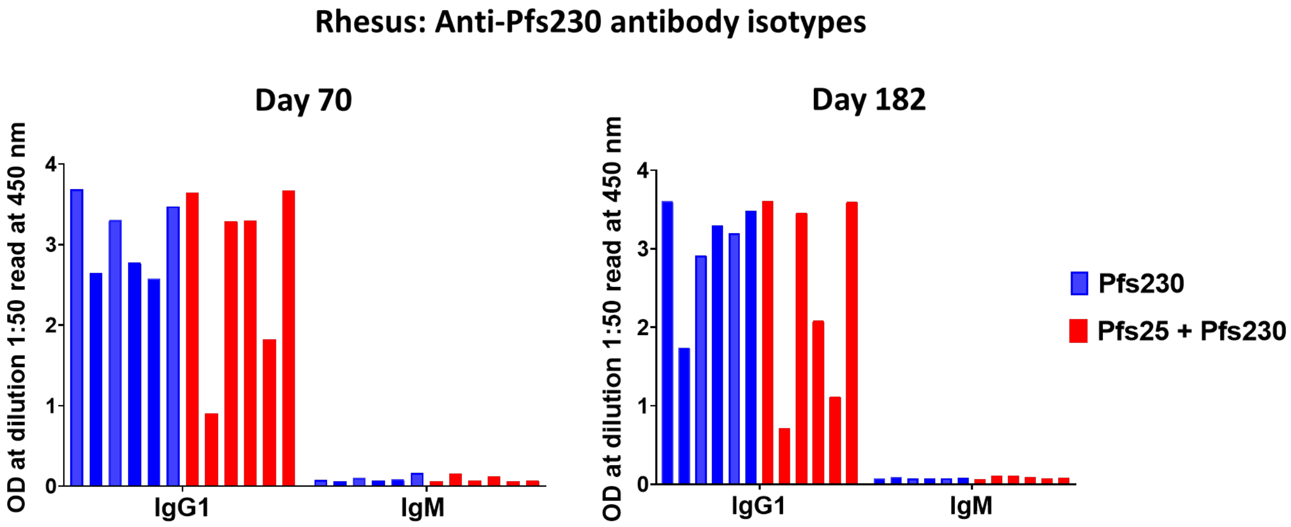

**Fig. S5. Isotyping of antibodies in sera from rhesus macaques that received either Pfs230D1-EPA/Alhydrogel® alone (n=6) or Pfs25-EPA/Alhydrogel® + Pfs230D1-EPA/Alhydrogel® combination (n=6).**

Antibody isotyping was performed on immune sera collected 2 weeks after the second (day 70) and third (day 182) vaccination dose by ELISA**.** OD values for IgG1and IgM ELISA assays are displayed for each individual animal that received either Pfs230D1-EPA/Alhydrogel® alone (Blue bars) or Pfs25-EPA/Alhydrogel® + Pfs230D1-EPA/Alhydrogel® combination (red bars).

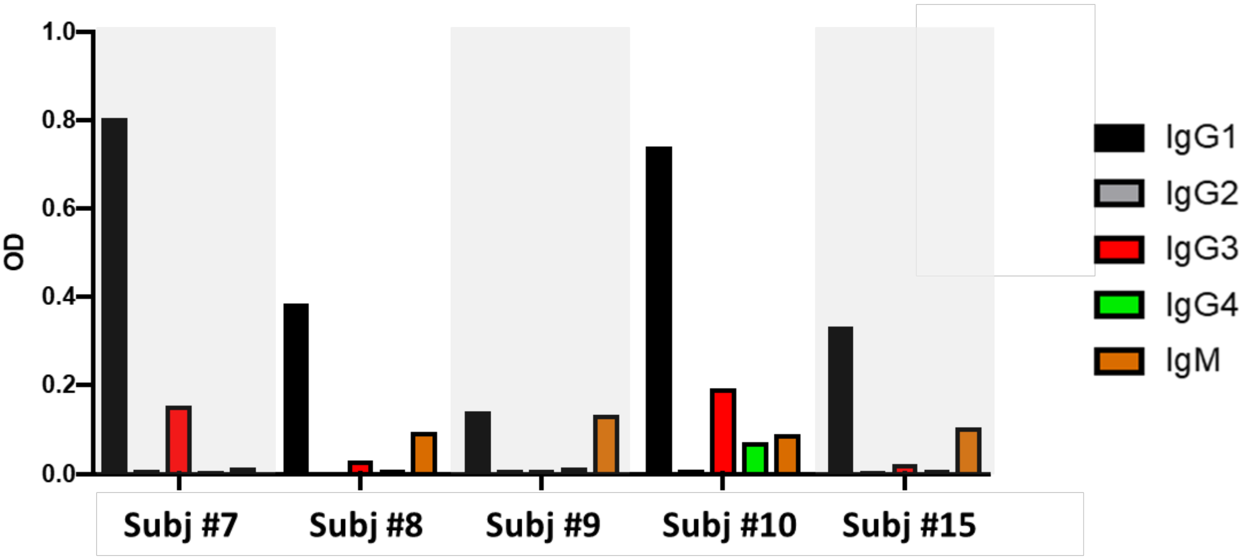

**Fig. S6. Isotyping of Pfs230D1-specific antibodies in sera from five US vaccinees with high antibody titers against Pfs230.**

ELISAs were performed on sera collected 2 weeks after the second vaccination. For each participant, individual ELISA OD values for IgG1, IgG2, IgG3, IgG4 and IgM are displayed.

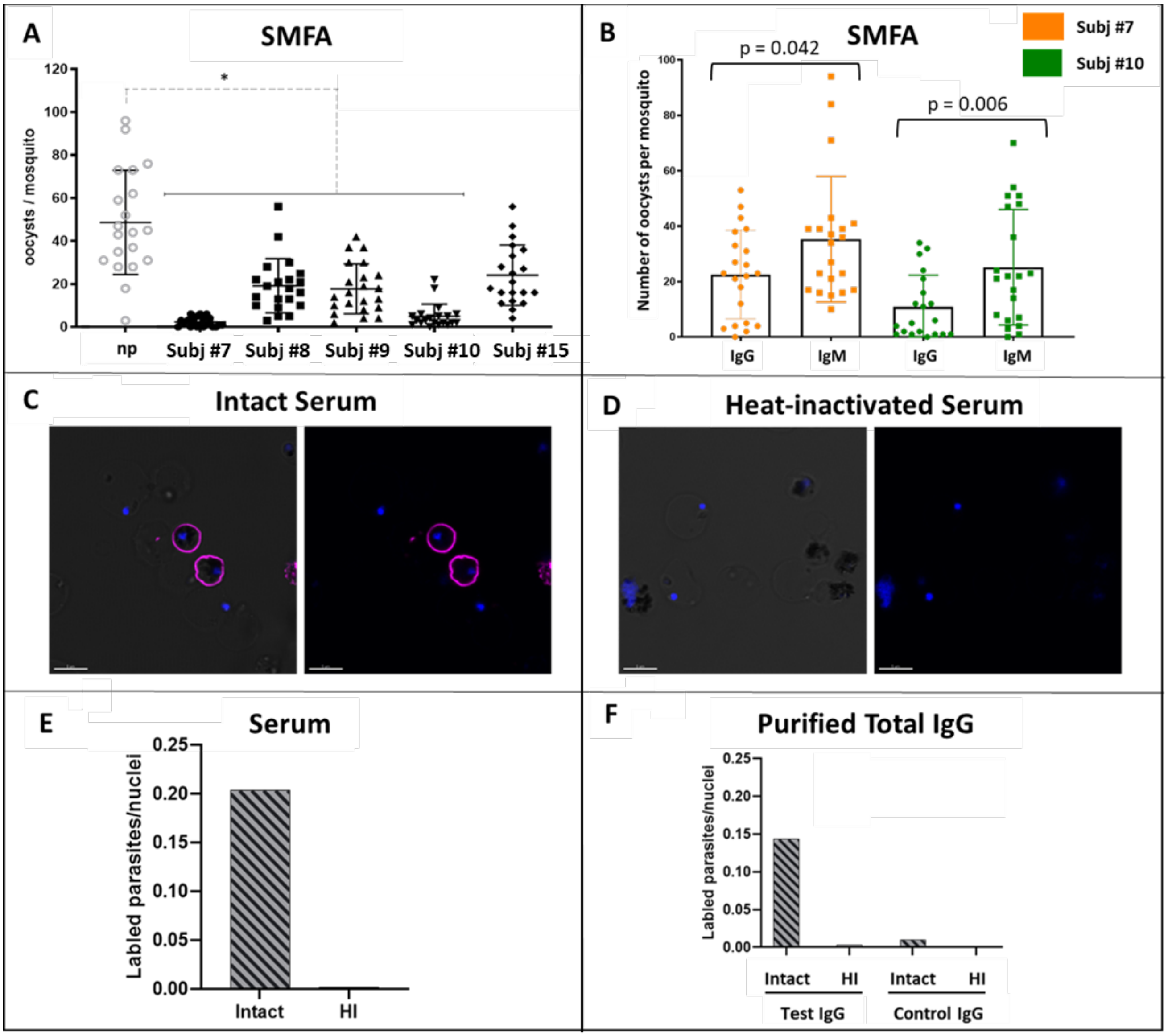

**Fig. S7. Functional activity of serum and purified IgG eight weeks post-vaccination (Day 84) including serum TRA and membrane attack complex formation on the parasite surface.**

(A) Sera collected 8 weeks post-second vaccination (Day 84) were analyzed by SMFA for 5 vaccinees with high TRA on Day 56. Differences between vaccinee sera and naïve pooled sera ("np") were analyzed by Kruskal-Wallis tests with Bonferroni correction for multiple comparisons; * indicates p value < 0.05. (B) IgG and IgM purified from Day 84 antisera of Subjects #7 and #10 were analyzed at neat concentration for comparative functional activity by SMFA, in the presence of intact non-immune sera. Differences between IgG and IgM were analyzed by a two sample Student's T test; p values for comparison within each subject are indicated in graph; when both subjects were combined, p = 0.001. Live IFA images captured by confocal microscopy of *P. falciparum* strain NF54 gametes incubated with intact (C) or heat-inactivated serum (D) from Subject #7 showing surface-deposited MAC (membrane attack complex) in presence of intact but not heat-inactivated serum. MAC was detected with Alexa 488-labeled antibody that recognizes the assembled MAC complex. Cell nuclei were labeled with Hoechst stain to differentiate parasites from contaminating red blood cells. The number of gametes showing MAC formation were quantified as a fraction of the total number of Hoechst-stained nuclei. MAC deposition was observed when gametes were incubated with either intact subject serum or purified IgG supplemented with naïve serum (C, E, F) and this deposition was lost after serum was heat-inactivated (D, E, F), since the heat-labile components of the complement pathway were degraded.

| mouse sera | immunogen | Vaccine Dose (µg/0.5 mL) | Serum dilution | anti-Pfs25 (EU) | anti-Pfs230D1 (EU) | mean oocysts/ mosquito | #infected/ #dissected |
|---|---|---|---|---|---|---|---|
| intact | Naïve | n/a | 1:8 | -- | -- | 39.5 | 23/23 |
| | Pfs25-EPA | 0.1 | 1:8 | 28,293 | | 0.7* | 7/23 |
| | | 0.1 | 1:32 | 7,073 | | 7.8* | 21/23 |
| | | 0.1 | 1:128 | 1,768 | | 27.1 | 24/24 |
| | Pfs230D1-EPA | 0.3 | 1:8 | | 4,085 | 12.5* | 21/24 |
| | | 0.3 | 1:32 | | 1,021 | 48.7 | 24/24 |
| | | 0.01 | 1:8 | | 390 | 41.5 | 23/23 |
| heat-inactivated | naive | n/a | 1:8 | -- | -- | 49.3 | 21/22 |
| | Pfs25-EPA | 0.1 | 1:8 | 28,293 | | 0.8* | 9/24 |
| | | 0.1 | 1:32 | 7,073 | | 3.8* | 17/24 |
| | | 0.1 | 1:128 | 1,768 | | 15.5* | 20/23 |
| | Pfs230D1-EPA | 0.3 | 1:8 | | 4,085 | 6.6* | 18/22 |
| | | 0.3 | 1:32 | | 1,021 | 16.8 | 20/23 |
| | | 0.01 | 1:8 | | 390 | 14.5* | 25/25 |

*p<0.05 by Kruskal-Wallis with Dunn's correction for multiple comparisons, when comparing post-vaccine to naïve sera

**Table S1: Sera from BALB/c mice immunized with Pfs25-EPA/Alhydrogel or Pfs230D1-EPA/Alhydrogel block infection in the presence or absence of complement.** Antibody titers by ELISA (anti-Pfs25; anti-Pfs230D1) and antibody function by SMFA are shown for sera taken before ("naïve") and after 2 vaccine doses. Three dilutions of sera from each group were used to titrate the activity. Titers shown are the amount of antibody in the mosquito feeder after dilution. EU = ELISA units. The average number of oocysts in mosquitoes fed with control human AB+ sera was 61. Table data are also represented in **Figure S3**.

| Vaccine Group | Animal | Titers in feeder (EU) | | mean oocysts/mosquito | | % TRA | Infected/Dissected | | % TBA |
|---|---|---|---|---|---|---|---|---|---|
| | | Pfs25 | Pfs230D1 | Pre (D0) | Post (D182) | | pre | post | |
| Pfs25-EPA/alum | 1 | 3,562 | | 18.1 | 10.5 | 42 | 21/24 | 17/23 | 16 |
| | 2 | 4,194 | | 13.7 | 6.4* | 53 | 22/24 | 22/24 | 0 |
| | 3 | 1,745 | | 19.5 | 26.4 | -35 | 19/23 | 22/24 | -11 |
| | 4 | 8,854 | | 48.3 | 11.6* | 76 | 23/23 | 22/23 | 4 |
| | 5 | 4,186 | | 29.7 | 27.7 | 7 | 22/22 | 24/24 | 0 |
| | 6 | 3,210 | | 42.1 | 15.2* | 64 | 19/23 | 18/20 | -9 |
| Pfs230D1-EPA/alum | 7 | | 13,499 | 9.4 | 0.1* | 99 | 20/20 | 2/26 | 92 |
| | 8 | | 4,123 | 22.0 | 2.2* | 90 | 26/26 | 21/26 | 19 |
| | 9 | | 7,249 | 19.0 | 0.4* | 98 | 26/26 | 8/25 | 68 |
| | 10 | | 4,601 | 52.2 | 1.6* | 97 | 21/21 | 15/22 | 32 |
| | 11 | | 3,449 | 68.4 | 3.6* | 95 | 22/22 | 18/22 | 18 |
| | 12 | | 8,916 | 64.4 | 0.8* | 99 | 20/20 | 7/20 | 65 |

*$p<0.05$ by Wilcoxon matched-pairs signed rank test, comparing D0 to D182

**Table S2: Rhesus anti-Pfs230D1 inhibits parasite transmission better than anti-Pfs25.** Rhesus sera from 2 weeks after the 3rd vaccination (D182) were tested for function by SMFA. 60 µL of serum from each animal was diluted with 100 µL of $AB^+$ human sera, mixed with 100 µL gametocyte culture, and fed to mosquitos. Oocysts were measured 8 days later. Titers shown are the amount of antibody in the mosquito feeder after diluting. EU = ELISA units. Transmission-reducing activity (%TRA) and transmission-blocking activity (%TBA) are relative to the pre-vaccination sera from the same animal. The average oocyst counts in negative controls (human AB+ sera) ranged from 16-64. Table data are also represented in **Fig. S4**.

| Animal | Intact sera | | | | Heat-inactivated sera | | | |
|---|---|---|---|---|---|---|---|---|
| | **mean oocysts/mosquito** | **% TRA** | **infected/ dissected** | **% TBA** | **mean oocysts/mosquito** | **% TRA** | **Infected/ dissected** | **% TBA** |
| Pre-bleed pool | 7.8 | | 22/23 | | 5.4 | | 24/24 | |
| 7 | 0.04 | 99.5 | 1/24 | 95.7 | 1.4 * | 74.4 | 17/24 | 29.2 |
| 8 | 1.7 | 78.6 | 19/24 | 17.4 | 3.0 | 44.2 | 19/24 | 20.8 |
| 9 | 0.1 | 98.4 | 3/24 | 87.0 | 0.5 | 90.7 | 7/24 | 70.8 |
| 10 | 0.1 | 98.4 | 2/24 | 91.3 | 1.5 * | 72.1 | 17/24 | 29.2 |
| 11 | 0.3 | 96.8 | 4/24 | 82.6 | 2.0 * | 62.0 | 15/24 | 37.5 |
| 12 | 0.04 | 99.5 | 1/24 | 95.7 | 1.3 * | 76.7 | 17/24 | 29.2 |

*p<0.05 by Wilcoxon matched-pairs signed rank test, comparing NORM to Heat-inactivated

**Table S3: Rhesus anti-Pfs230D1 requires complement for optimal activity.**
Rhesus sera from 2 weeks after the 3rd vaccination (D182) of Pfs230D1-EPA were tested for function by SMFA. Sixty microliters (60 µL) of test sera was diluted with 100 µL of $AB^+$ human sera, intact (thus with complement) or heat-inactivated (thus without complement), mixed with 100 µL gametocyte culture, and fed to mosquitos. Oocysts were measured 8 days later. Transmission-reducing activity (%TRA) and transmission-blocking activity (%TBA) are relative to the pre-bleed pools. As a group, serum TRA was significantly greater in intact versus heat-inactivated sera (P<0.05, Wilcoxon 2-tailed signed-rank test). The average oocyst counts in negative controls (human AB+ sera) ranged from 16-64.

| Pfs25M-EPA/Alhydrogel® Alone | | | | | | |
|---|---|---|---|---|---|---|
| | **Pfs25M, 16µg** | | | **Pfs25M, 47µg** | | |
| | **Vaccine 1** | **Vaccine 2** | **Total** | **Vaccine 1** | **Vaccine 2** | **Total** |
| | N=5 | N=4 | N=5 | N=5 | N=5 | N=5 |
| Total # AEs | 17 (3) 60% | 10 (2) 50% | 27 (4) 80% | 11 (5) 100% | 13 (5) 100% | 24 (5) 100% |
| **Classification** | | | | | | |
| Local Reactogenicity | 4 (2) 40% | 2 (2) 50% | 6 (3) 60% | 6 (5) 100% | 7 (4) 80% | 13 (5) 100% |
| Systemic Reactogenicity | 8 (2) 40% | 0 (0) 0% | 8 (2) 40% | 1 (1) 20% | 0 (0) 0% | 1 (1) 20% |
| Laboratory Abnormalities | 3 (2) 40% | 1 (1) 25% | 4 (3) 60% | 2 (2) 40% | 2 (2) 40% | 4 (3) 60% |
| Unsolicited AEs | 2 (2) 40% | 7 (1) 25% | 9 (3) 60% | 2 (2) 40% | 4 (3) 60% | 6 (5) 100% |
| **Severity and Relationship** | | | | | | |
| Grade 1 | 13 (2) 40% | 10 (2) 50% | 23 (4) 80% | 11 (5) 100% | 10 (4) 80% | 21 (5) 100% |
| *Pfs25 Related* | 9 (2) 40% | 2 (2) 50% | 11 (3) 60% | 8 (5) 100% | 6 (4) 80% | 14 (5) 100% |
| Grade 2 | 4 (2) 40% | 0 (0) 0% | 4 (2) 40% | 0 (0) 0% | 1 (1) 20% | 1 (1) 20% |
| *Pfs25 Related* | 2 (2) 40% | 0 (0) 0% | 2 (2) 40% | 0 (0) 0% | 0 (0) 0% | 0 (0) 0% |
| Grade 3 | 0 (0) 0% | 0 (0) 0% | 0 (0) 0% | 0 (0) 0% | 2 (2) 40% | 2 (2) 40% |
| *Pfs25 Related* | 0 (0) 0% | 0 (0) 0% | 0 (0) 0% | 0 (0) 0% | 0 (0) 0% | 0 (0) 0% |
| Grade 4 | 0 (0) 0% | 0 (0) 0% | 0 (0) 0% | 0 (0) 0% | 0 (0) 0% | 0 (0) 0% |
| SAE | 0 (0) 0% | 0 (0) 0% | 0 (0) 0% | 0 (0) 0% | 0 (0) 0% | 0 (0) 0% |

| Pfs230D1-EPA/Alhydrogel® Alone | | | | | | | | | |
|---|---|---|---|---|---|---|---|---|---|
| | **Pfs230D1, 5μg** | | | **Pfs230D1, 15μg** | | | **Pfs230D1, 40μg** | | |
| | **Vaccine 1** | **Vaccine 2** | **Total** | **Vaccine 1** | **Vaccine 2** | **Total** | **Vaccine 1** | **Vaccine 2** | **Total** |
| | N=5 | N=5 | N=5 | N=5 | N=4 | N=5 | N=5 | N=5 | N=5 |
| Total # AEs | 8 (4) 80% | 14 (5) 100% | 22 (5) 100% | 10 (5) 100% | 24 (4) 100% | 34 (5) 100% | 19 (5) 100% | 12 (5) 100% | 31 (5) 100% |
| **Classification** | | | | | | | | | |
| Local Reactogenicity | 2 (2) 40% | 4 (4) 80% | 6 (4) 80% | 5 (4) 80% | 4 (3) 75% | 9 (4) 80% | 7 (5) 100% | 4 (4) 80% | 11 (5) 100% |
| Systemic Reactogenicity | 1 (1) 20% | 1 (1) 20% | 2 (2) 40% | 3 (2) 40% | 6 (2) 50% | 9 (3) 60% | 6 (3) 60% | 0 (0) 0% | 6 (3) 60% |
| Laboratory Abnormalities | 0 (0) 0% | 2 (2) 40% | 2 (2) 40% | 1 (1) 20% | 0 (0) 0% | 1 (1) 20% | 3 (1) 20% | 1 (1) 20% | 4 (2) 40% |
| Unsolicited AEs | 5 (3) 60% | 7 (2) 40% | 12 (3) 60% | 1 (1) 20% | 14 (4) 100% | 15 (4) 80% | 3 (2) 40% | 7 (5) 100% | 10 (5) 100% |
| **Severity and Relationship** | | | | | | | | | |
| Grade 1 | 6 (4) 80% | 13 (5) 100% | 19 (5) 100% | 10 (5) 100% | 19 (4) 100% | 29 (5) 100% | 15 (5) 100% | 9 (5) 100% | 24 (5) 100% |
| *Pfs230 Related* | 3 (3) 60% | 6 (4) 80% | 9 (4) 80% | 7 (4) 80% | 9 (3) 75% | 16 (4) 80% | 9 (5) 100% | 5 (4) 80% | 14 (5) 100% |
| Grade 2 | 2 (2) 40% | 1 (1) 20% | 3 (3) 60% | 0 (0) 0% | 5 (3) 75% | 5 (3) 60% | 4 (2) 60% | 3 (3) 60% | 7 (4) 80% |

| | | | | | | | | | |
|---|---|---|---|---|---|---|---|---|---|
| *Pfs230 Related* | 0 (0) 0% | 0 (0) 0% | 0 (0) 0% | 0 (0) 0% | 0 (0) 0% | 0 (0) 0% | 1 (1) 20% | 0(0) 0% | 1 (1) 20% |
| Grade 3 | 0 (0) 0% | 0 (0) 0% | 0 (0) 0% | 0 (0) 0% | 0 (0) 0% | 0 (0) 0% | 0 (0) 0% | 0 (0) 0% | 0 (0) 0% |
| Grade 4 | 0 (0) 0% | 0 (0) 0% | 0 (0) 0% | 0 (0) 0% | 0 (0) 0% | 0 (0) 0% | 0 (0) 0% | 0 (0) 0% | 0 (0) 0% |
| SAE | 0 (0) 0% | 0 (0) 0% | 0 (0) 0% | 0 (0) 0% | 0 (0) 0% | 0 (0) 0% | 0 (0) 0% | 0 (0) 0% | 0 (0) 0% |

| Pfs25M-EPA/Alhydrogel® + Pfs230D1-EPA/Alhydrogel® | | | | | | |
|---|---|---|---|---|---|---|
| | Pfs25M, 16µg AND Pfs230D1, 15µg | | | Pfs25M, 47µg AND Pfs230D1, 40µg | | |
| | **Vaccine 1** | **Vaccine 2** | **Total** | **Vaccine 1** | **Vaccine 2** | **Total** |
| | N=5 | N=5 | N=5 | N=5 | N=5 | N=5 |
| Total # AEs | 14 (5) 100% | 13 (5) 100% | 27 (5) 100% | 23 (5) 100% | 21 (5) 100% | 44 (5) 100% |
| **Classification** | | | | | | |
| Local Reactogenicity | 6 (4) 80% | 8 (4) 80% | 14 (4) 80% | 9 (5) 100% | 11 (4) 80% | 20 (5) 100% |
| Systemic Reactogenicity | 3 (2) 40% | 1 (1) 20% | 4 (2) 40% | 5 (2) 40% | 4 (1) 20% | 9 (2) 40% |
| Laboratory Abnormalities | 2 (2) 40% | 2 (1) 20% | 4 (2) 40% | 1 (1) 20% | 0 (0) 0% | 1 (1) 20% |
| Unsolicited AEs | 3 (3) 60% | 2 (2) 40% | 5 (4) 80% | 8 (4) 80% | 6 (2) 40% | 14 (5) 100% |
| **Severity and Relationship** | | | | | | |
| Grade 1 | 14 (5) 100% | 13 (5) 100% | 27 (5) 100% | 22 (5) 100% | 20 (5) 100% | 42 (5) 100% |
| *Related to Pfs25* | 4 (2) 40% | 4 (4) 80% | 8 (4) 80% | 9 (5) 100% | 7 (3) 60% | 16 (5) 100% |
| *Related to Pfs230* | 6 (4) 80% | 4 (3) 80% | 10 (4) 80% | 9 (5) 100% | 8 (4) 80% | 17 (5) 100% |
| Grade 2 | 0 (0) 0% | 0 (0) 0% | 0 (0) 0% | 0 (0) 0% | 1 (1) 20% | 1 (1) 20% |
| *Related to Pfs25* | 0 (0) 0% | 0 (0) 0% | 0 (0) 0% | 0 (0) 0% | 0 (0) 0% | 0 (0) 0% |
| *Related to Pfs230* | 0 (0) 0% | 0 (0) 0% | 0 (0) 0% | 0 (0) 0% | 0 (0) 0% | 0 (0) 0% |
| Grade 3 | 0 (0) 0% | 0 (0) 0% | 0 (0) 0% | 1 (1) 20% | 0 (0) 0% | 1 (1) 20% |
| *Related to Pfs25* | 0 (0) 0% | 0 (0) 0% | 0 (0) 0% | 0 (0) 0% | 0 (0) 0% | 0 (0) 0% |
| *Related to Pfs230* | 0 (0) 0% | 0 (0) 0% | 0 (0) 0% | 0 (0) 0% | 0 (0) 0% | 0 (0) 0% |
| Grade 4 | 0 (0) 0% | 0 (0) 0% | 0 (0) 0% | 0 (0) 0% | 0 (0) 0% | 0 (0) 0% |
| SAE | 0 (0) 0% | 0 (0) 0% | 0 (0) 0% | 0 (0) 0% | 0 (0) 0% | 0 (0) 0% |

**Table S4. Summary of adverse events from human clinical trial evaluating the safety of Pfs25M-EPA/ Alhydrogel® versus Pfs230D1-EPA/Alhydrogel® versus combination of the two.**

Local injection site reactions (including pain/tenderness, erythema/redness, swelling, induration, and pruritus) were assessed until day 7 after vaccination or until resolved. All systemic solicited reactogenicity (including fever, headache, nausea, malaise, myalgia, arthralgia, and urticaria) and unsolicited AEs were recorded through day 14 after each vaccination. Both solicited local and systemic reactogenicity were solicited from the subjects during clinic visits and with daily diary cards through day 7 post vaccination. Similar to solicited AEs, all laboratory AEs were collected and graded through 14 days after each vaccination or until resolved. X(X)X% = absolute number of AE (number of subjects experiencing AEs) percentage of subjects with AEs. SAE = serious adverse events.

| Vaccine arm | subject | anti-Pfs25 (EU) | anti-Pfs230D1 (EU) | mean oocysts/mosquito | %TRA | infected/dissected | %TBA |
|---|---|---|---|---|---|---|---|
| Pfs25-EPA | 1 | 158 | | 49.8 | -24 | 23/24 | 4 |
| | 2 | 453 | | 47.8 | -19 | 24/24 | 0 |
| | 3 | 686 | | 30.8 | 23 | 23/24 | 4 |
| | 4 | 17 | | 52.4 | -31 | 24/24 | 0 |
| | 5 | 126 | | 43.9 | -10 | 24/24 | 0 |
| Pf230D1-EPA | 6 | | 32 | 52.9 | -32 | 24/24 | 0 |
| | 7 | | 741 | 0.04 * | 99 | 1/25*** | 96 |
| | 8 | | 204 | 16.5 * | 59 | 21/24 | 13 |
| | 9 | | 77 | 19.0 | 52 | 23/24 | 4 |
| | 10 | | 512 | 0.9 * | 98 | 13/25*** | 48 |
| Pfs25-EPA + Pfs230D1-EPA | 11 | 17 | 23 | 49.4 | -23 | 24/24 | 0 |
| | 12 | 28 | 44 | 59.3 | -48 | 24/24 | 0 |
| | 13 | 24 | 93 | 46.7 | -17 | 24/24 | 0 |
| | 14 | 64 | 418 | 14.1 * | 65 | 23/24 | 4 |
| | 15 | 122 | 198 | 4.1 * | 90 | 19/25** | 24 |
| pre-bleed pool | - | - | - | 40.0 | - | 24/24 | - |

* $p<0.05$, Kruskal-Wallis with Dunn's correction for multiple comparison; **$p<0.05$, ***$p<0.001$, Fisher exact test

**Table S5: Anti-Pfs230D1 has functional activity in humans after 2 doses.**
Sera collected 2 weeks after the 2nd vaccination were tested for function by SMFA. 160 µL of sera was mixed with 100 µL gametocyte culture and fed to mosquitos. Oocysts were measured 8 days later. Transmission-reducing activity (%TRA) and transmission-blocking activity (%TBA) are relative to the pre-vaccination pool. Table data are also represented in **Fig. 3**.

| | | Intact sera | | | Heat-inactivated sera | | |
|---|---|---|---|---|---|---|---|
| | | **mean oocysts/mosquito** | | **% TRA** | **mean oocysts/mosquito** | | **% TRA** |
| | **subject** | **pre-bleed** | **D42** | | **pre-bleed** | **D42** | |
| Pfs230D1-EPA | 6 | 17.3 | 18.0 | -4 | 12.6 | 11.6 | 7.6 |
| | 7 | 12.1 | 0 * | 100 | 11.5 | 5.8 | 49.5 |
| | 8 | 17.4 | 6.6 * | 61.8 | 12.3 | 10.3 | 15.9 |
| | 9 | 25.8 | 6.8 * | 73.6 | 13.5 | 17.4 | -29.4 |
| | 10 | 20.2 | 0.3 * | 98.4 | 18.8 | 5.6 * | 70.1 |
| Pfs25-EPA + Pfs230D1-EPA | 11 | 25.3 | 13.7 | 45.8 | 12.9 | 32.9 | -155.0 |
| | 12 | 22.5 | 29.8 | -32.4 | 15.8 | 31.9 | -101.9 |
| | 13 | 23.2 | 23.5 | -1.6 | 27.6 | 34.3 | -24.2 |
| | 14 | 11.2 | 3.4 * | 69.9 | 12.2 | 12.9 | -5.7 |
| | 15 | 20.7 | 1.9 * | 90.7 | 17.5 | 16.2 | 8.5 |

*P<0.05, Wilcoxon matched-pairs signed rank test

**Table S6: Anti-Pfs230D1 requires complement for activity in humans.**
Sera from 2 weeks after the 2nd vaccination of Pfs230D1-EPA or Pfs230D1-EPA+Pfs25-EPA were tested for function by SMFA. Half of each sample was heat-treated, and 160 µL of sera was mixed with 100 µL gametocyte culture and fed to mosquitos. Oocysts were measured 8 days later. %TRA and %TBA are relative to the pre-vaccination sera. Table data are also represented in **Fig. 4**. The average oocyst counts in negative controls (human AB+ sera) was 14.

| Target Species | Antibody isotype | Detecting antibody clone | Supplier |
|---|---|---|---|
| Human | IgG1 | HP6069 | Invitrogen |
| Human | IgG2 | HP6014 | Invitrogen |
| Human | IgG3 | HP6047 | Invitrogen |
| Human | IgG4 | HP6025 | Invitrogen |
| Human | IgM | HP6083 | Invitrogen |
| Rhesus | IgG1 | 7H11 | Nonhuman primate reagent resource |
| Rhesus | IgG2 | 3C10 | Nonhuman primate reagent resource |
| Rhesus | IgG3 | 2G11 | Nonhuman primate reagent resource |
| Rhesus | IgM | Polyclonal | Jackson Immunoresearch Inc. |

**Table S7: Detecting antibodies used for Pfs230 isotyping assays.**
The list of detecting antibodies that were used to enumerate specific isotypes against Pfs230 in human and rhesus samples after vaccinations are displayed. 3C10 and 2G11 monoclonals for detection of IgG2 and IgG3 in rhesus could not be validated.
